# Association of Topiramate Prescribed for any Indication with Reduced Alcohol Consumption in Electronic Health Record Data

**DOI:** 10.1101/2022.05.17.22275219

**Authors:** Henry R. Kranzler, Shirley H. Leong, Michelle Naps, Emily E. Hartwell, David A. Fiellin, Christopher T. Rentsch

## Abstract

**Background and Aims:** Topiramate is widely prescribed to treat a variety of conditions, including alcohol use disorder (AUD). We used electronic health record (EHR) data to examine topiramate’s effects on drinking in individuals differentiated by a history of AUD.

**Design:** Parallel-groups comparison of patients prescribed topiramate and a propensity-score matched comparison group.

**Setting:** A large U.S. integrated healthcare system.

**Participants:** Patients with Alcohol Use Disorders Identification Test-Consumption (AUDIT-C) scores prior to and after a minimum of 180 days of topiramate prescription for any indication and a propensity-score matched group. The sample included 5,918 patients with an electronic health record diagnosis of alcohol use disorder at any time (AUD-hx-pos) (1,738 topiramate exposed and 4,180 controls) and 23,614 patients with no EHR diagnosis of AUD (AUD-hx-neg) (6,324 topiramate exposed and 17,290 controls).

**Measurements:** Regression analyses compared difference-in-difference (DiD) estimates, separately by AUD history. DiD estimates represent exposure-group (i.e., topiramate vs. control) differences on the pre-post difference in AUDIT-C score. Effects of baseline AUDIT-C score and daily topiramate dosage were also tested.

**Findings:** Among AUD-hx-neg patients, those who received topiramate had a greater reduction in AUDIT-C score (−0.11) than matched controls (−0.04). This yielded a DiD score of -0.07 (95% CI= -0.11,-0.03; *P* =0.002), with the greatest effect among AUD-hx-neg patients with a baseline AUDIT-C score of 4+ (DiD = -0.35, 95% CI=-0.49, -0.21; *P* <0.0001) and those prescribed >150 mg/day of the medication (DiD = -0.15, 95%CI=-0.23, -0.07; *P* <0.001).

**Discussion:** The lack of an effect of topiramate on drinking levels in AUD-hx-pos patients contrasts with the robust reductions seen in topiramate clinical trials. Research is needed to ascertain whether AUDIT-C scores from EHR data accurately reflect medication effects on drinking and whether patient characteristics can be used to select patients most likely to reduce their drinking when treated with topiramate.

## INTRODUCTION

Topiramate, first approved by the U.S. Food and Drug Administration (FDA) in 1996 as an anticonvulsant, was subsequently approved to prevent migraine and, in combination with phentermine, for weight loss. It is widely prescribed for all three indications (1-4), with 7.5 to 10.75 million U.S. topiramate prescriptions annually from 2010 to 2018 (5). Although topiramate is not approved for treating alcohol use disorder (AUD), based on randomized controlled clinical trials showing that the medication is efficacious in promoting abstinence and reducing heavy drinking and alcohol craving (6), it is now considered an evidence-based treatment for moderate-to-severe AUD (7-9).

Given these findings, topiramate could reduce drinking among individuals who receive it for an indication other than treating AUD. Further, the effect of topiramate on alcohol use could differ among individuals depending on whether they had ever been diagnosed with AUD and as a function of either baseline drinking level or average daily dosage of the drug. To address these questions, we used data from the largest integrated healthcare system in the United States to examine topiramate’s effects separately for individuals known to have a history of AUD (AUD-hx-pos) and those without evidence of such a history (AUD-hx-neg). We anticipated that topiramate-treated patients would show a greater reduction in AUDIT-C scores than an unexposed control group among both AUD-hx-pos and AUD-hx-neg patients. We also expected that the effects of topiramate would increase with increasing pretreatment AUDIT-C scores and an increasing average daily topiramate dosage.

## METHODS

### Study population

We performed an observational cohort study using data from the US Department of Veterans Affairs (VA) Corporate Data Warehouse (CDW), comprising the electronic health records (EHR) of >17 million patients accessing care at more than 1,200 U.S. hospitals, medical centers, and community outpatient clinics (10). Available data include information on all outpatient and inpatient encounters, including demographics, medical and psychiatric diagnoses, pharmacy dispensing records, laboratory measures, and routinely collected measures of smoking and alcohol consumption.

We identified a cohort of 1,044,029 patients who were dispensed topiramate at VA pharmacies between January 1, 2009 and October 1, 2015 (to ensure consistency in disease coding as that was when ICD-10 was implemented system-wide to replace ICD-9) or matched controls with no history of topiramate exposure. The topiramate-exposed group included all patients who received the medication for at least 180 continuous days for any indication, with coverage for at least 144 of those days and no gap larger than 30 days. We identified new episodes of topiramate exposure by applying a washout period, so that patients who received topiramate at any point were eligible to be followed only after one year with no exposure. The clinics that commonly generated topiramate prescriptions included: primary care, neurology, psychiatry, women’s health, substance use disorder treatment, pain medicine, endocrinology, physical medicine and rehabilitation, and clinical pharmacy. For unexposed controls, we selected patients who attended at least one of these clinics but never received topiramate to ensure that unexposed patients came from the same source population and had an equal opportunity to receive topiramate. We defined the index date as the first fill date for topiramate-exposed patients and a random outpatient visit date per calendar year for unexposed patients.

We excluded patients with no outpatient care in the VA in the year prior to their index date, those with no AUDIT-C score, a routinely collected measure of alcohol consumption (11) in the two years prior to their index date, and those who had received treatment with medications that could reduce alcohol consumption. As can be seen in Supplementary Table 1, within both the AUD-hx-pos patients and the AUD-hx-neg patients, the prevalence of co-occurring psychiatric and substance use disorders did not differ as a function of topiramate treatment history. However, for virtually all co-occurring disorders, the AUD-hx-pos patients have a much higher prevalence of co-occurring disorders than do the AUD-hx-neg patients. Although topiramate is not approved to treat any psychiatric disorder, it is frequently prescribed off-label to treat mood disorders and substance use disorders, including AUD.

### Measures

An AUD history required the presence of one inpatient or two outpatient ICD-9 (303.X or 305-305.03) diagnostic codes at any time prior to baseline. A current AUD diagnosis had to be present during the year prior to baseline. Alcohol consumption was assessed using the AUDIT-C, a three-item, self-report screening questionnaire that detects hazardous or harmful drinking by querying past-year: 1) frequency of drinking, 2) number of drinks consumed on a typical day, and 3) frequency of drinking 6 or more drinks on an occasion (i.e., binge drinking) (11). AUDIT-C scores range from 0-12, with higher scores indicating greater risk of alcohol-related adverse effects. An AUDIT-C score of zero is defined as no current alcohol use, 1-3 indicates lower-risk drinking, 4-7 is considered at-risk drinking, and ≥8 is considered hazardous or harmful alcohol consumption. Since 2007, the VA has required annual AUDIT-C screening of all patients in primary care (12).

### Propensity-score matching

We used propensity-score matching on a set of covariates associated with the receipt of topiramate to adjust for the conditional probability of being prescribed the medication. This reduced the risk that patients with specific alcohol consumption patterns would be more likely to receive topiramate (13). By balancing the exposed and unexposed groups, the design simulates the effect of treatment allocation in a randomized controlled trial (14). Because estimating propensity scores separately has been shown to be unbiased, particularly in subgroup analyses in small samples (15-17), we used multivariable logistic regression models to estimate propensity scores separately for AUD-hx-pos and AUD-hx-neg patients, as defined below.

Variables used in the propensity-score models were selected *a priori* based on clinical knowledge (18). They included the year of index date, age at baseline, race, smoking status, body mass index at baseline, site prescribing pattern (the proportion of patients who initiated topiramate stratified by year), hepatitis C virus status, history of seizure prior to baseline, history of pain diagnoses prior to baseline (including neuropathy, osteoarthritis, or pain in the abdomen, back, chest, extremity, or neck, headache, or fracture), and history of medical and psychiatric conditions prior to baseline (including atrial fibrillation, myocardial infarction/coronary artery disease, peripheral vascular disease, diabetes, nephrolithiasis, glomerulonephritis, hyperlipidemia, pancreatitis, drug use disorders, post-traumatic stress disorder (PTSD), major or other depression, anxiety disorder, bipolar disorder, schizophrenia and schizoaffective disorder). We also included variables that captured attendance at clinics (including primary care, dialysis, diabetic retinal screening, rheumatology, infectious disease, nephrology, neurology, pain, allergy, chiropractic, dental, diabetes, emergency department, electrocardiogram lab, ophthalmology, hematology, oncology, homeless program, nutrition, orthopedics, substance use, mental health, PTSD), frequency of all-cause hospitalizations, and the total number of unique clinics visited in the year prior to baseline. Lastly, variables denoting receipt of other prescriptions (starting on or before the index date and continuing past the index date) to treat pain (including non-steroidal anti-inflammatory drugs, opioids, muscle relaxants, and antidepressants) were included in the model. Significant interaction terms (*P* < 0.05) were kept in the final model. The model C-statistic was 0.846 for patients with AUD and 0.886 for patients without AUD, indicating adequate discrimination between topiramate-exposed and -unexposed patients in both models (19).

Because the distribution of propensity scores for exposed and unexposed patients differed, we used propensity-score matching to exclude non-exchangeable unexposed patients (20). We conducted propensity-score matching within pre-specified subgroups of patients based on baseline AUDIT-C score (four groups: 0, 1-3, 4-7, 8+) and year of the index date (each year from 2009 through 2015) and aggregated the strata to create the matched cohort (21). Each exposed patient was matched to up to five unexposed patients with index dates in the same calendar year, using a greedy matching algorithm (22).

### Follow-up period

We followed patients from their index date for a maximum of two years or until their last VA visit or death. Additionally, topiramate-exposed patients were censored at 30 days after the end of their last topiramate prescription (allowing for a maximum 30-day gap between fills). To ensure equal follow-up time within matched sets, unexposed patients were censored at the total follow-up time of their matched exposed patient. Once patients were censored, their AUDIT-C scores were not included in the analyses.

### Statistical analyses

Statistical analyses were performed separately for patients with an AUD history (AUD-hx-pos) and those with no AUD diagnosis code (AUD-hx-neg) as of their index date. Although the availability of an AUDIT-C score was a criterion for study inclusion, we did not restrict matching eligibility to patients with a follow-up AUDIT-C score (the outcome measure), as no such exclusion would be applied in a randomized clinical trial. To maintain a balanced sample, for all exposed patients with no follow-up AUDIT-C, we removed the entire set of matched, unexposed patients. If an unexposed patient had no follow-up AUDIT-C score, we excluded only that individual, retaining all other unexposed patients in the set. We used the phi coefficient (>0.05) and the standardized mean difference (where SMD>0.1 indicates imbalance) to examine balance between exposed and unexposed patients in the full sample, the propensity-score matched sample, and the final analytic sample (23).

Among patients in the final analytic sample, we calculated the average baseline and follow-up AUDIT-C scores. The baseline AUDIT-C scores was defined as that closest to the index date, within a maximum of two years. Follow-up AUDIT-C scores were defined as the measure on the date closest to the end of the exposure and no later than 30 days post-exposure. We used multivariable difference-in-difference (DiD) linear regression models (24, 25) via repeated measures to estimate the differential change between baseline and follow-up AUDIT-C scores. Models included a binary indicator for treatment exposure, a binary indicator for time (baseline versus follow-up), and an exposure by time interaction term. All interactions were tested for significance with Type 3 tests of fixed effects. DiD estimates for the exposure groups were compared using t-tests. To account for residual confounding after propensity-score matching, models were adjusted for characteristics that differed between the exposed and unexposed groups post propensity-score matching. We used a phi coefficient>0.05 and SMD>0.1 to identify confounding categorical and continuous confounders, respectively.

The moderating effects of self-reported level of alcohol consumption at baseline and average daily topiramate dosage during the observation period were examined in three-way interactions with treatment exposure and time, with significance determined by Type 3 tests of fixed effects. We also performed subgroup analyses within significant moderators using Bonferroni-corrected t-tests on contrast estimates. Daily dosage was categorized into three levels of potential clinical relevance: low [≤100 milligrams (mg)], medium (>100 mg to 150 mg), and high (>150 mg). All statistical analyses were performed using SAS Enterprise Guide version 8.2 (SAS Institute Inc., Cary, NC, USA).

To test the sensitivity of the results to the exclusion of non-drinkers, we repeated the analyses after removing patients who reported no alcohol consumption as measured closest in time to the baseline assessment.

## RESULTS

### Study Sample

As shown in Figure 1, the initial sample comprised 1,044,029 patients, including 38,366 topiramate-exposed patients and 1,005,663 potential controls. Propensity-score matching yielded 22,332 AUD-hx-pos patients and 77,478 AUD-hx-neg patients. The application of exclusion criteria (i.e., patients receiving an exclusionary medication or lost to follow-up, or unexposed patients matched to exposed patients who were excluded or lost to follow-up) led to the exclusion of 16,414 AUD-hx-pos patients and 53,864 AUD-hx-neg patients. The final sample available for analysis consisted of 5,918 AUD-hx-pos patients (1,738 topiramate exposed and 4,180 controls) and 23,614 AUD-hx-neg patients (6,324 topiramate exposed and 17,290 controls).

**Figure 1:**
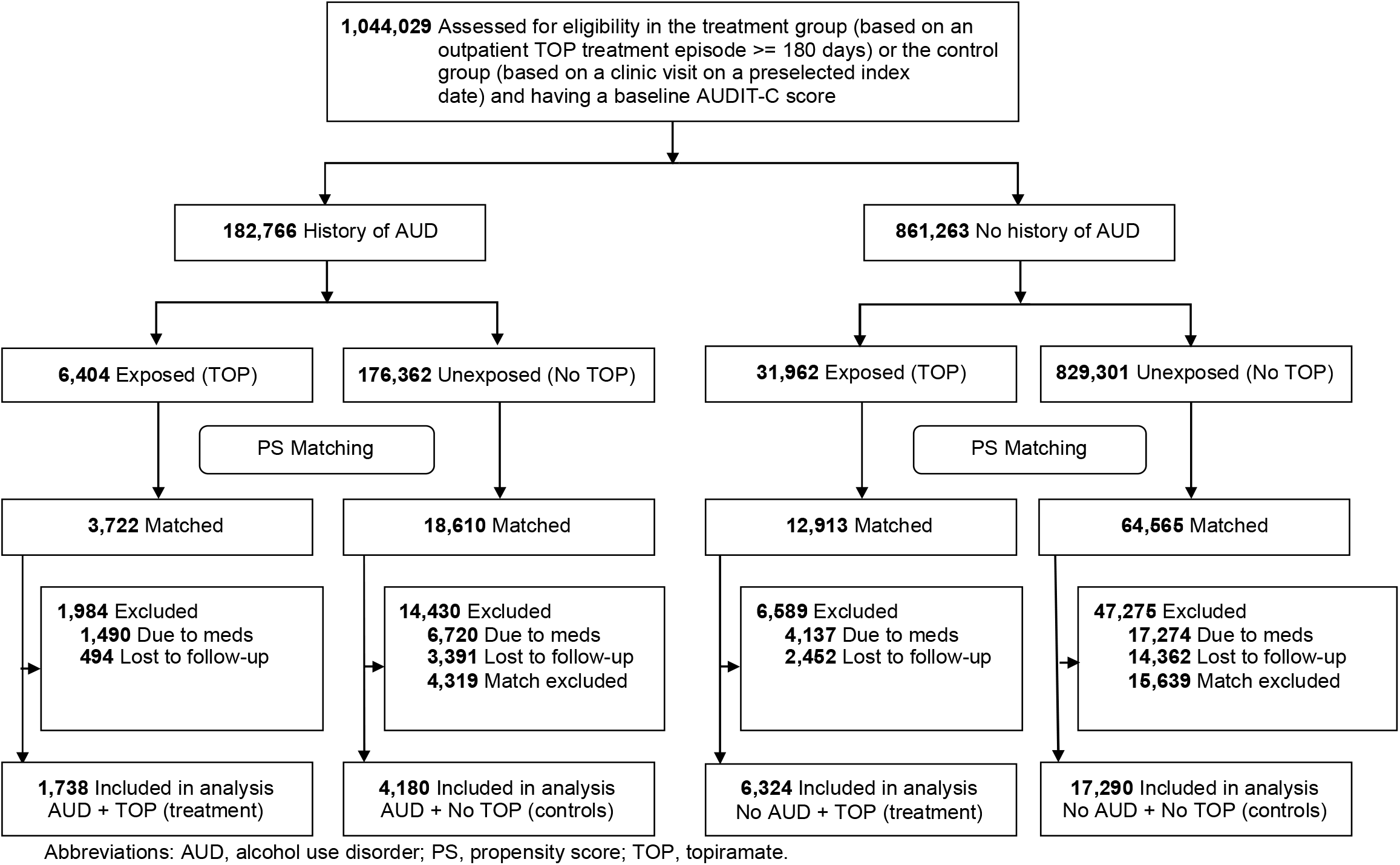
Selection of the Study Sample: Flow chart showing the numbers of patients retained for analysis following the application of inclusion and exclusion criteria

**Figure 2:**
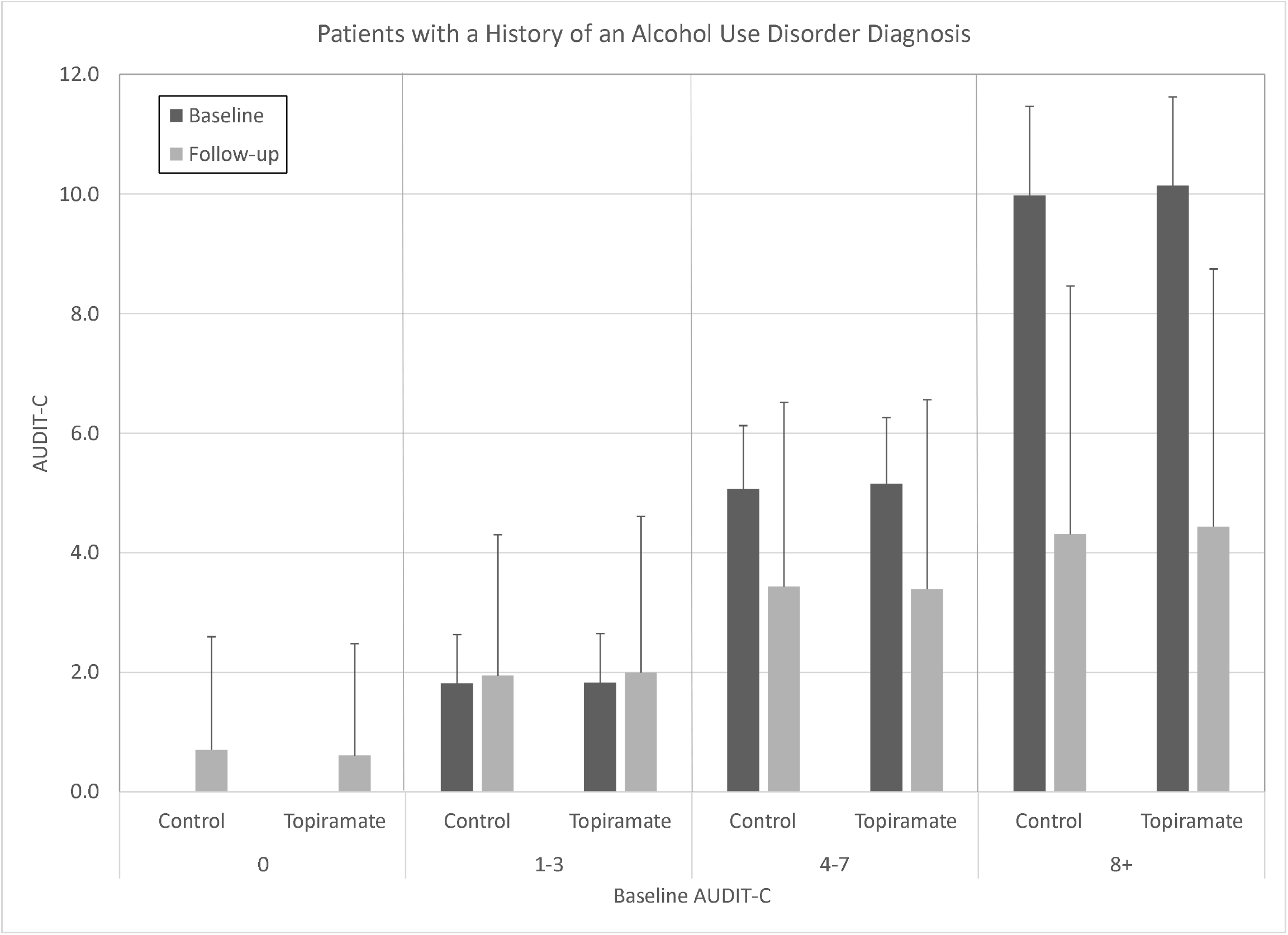
Baseline and Follow-up AUDIT-C Scores by Baseline AUDIT-C Score Group Among Patients with a History of an Alcohol Use Disorder Diagnosis.

**Figure 3:**
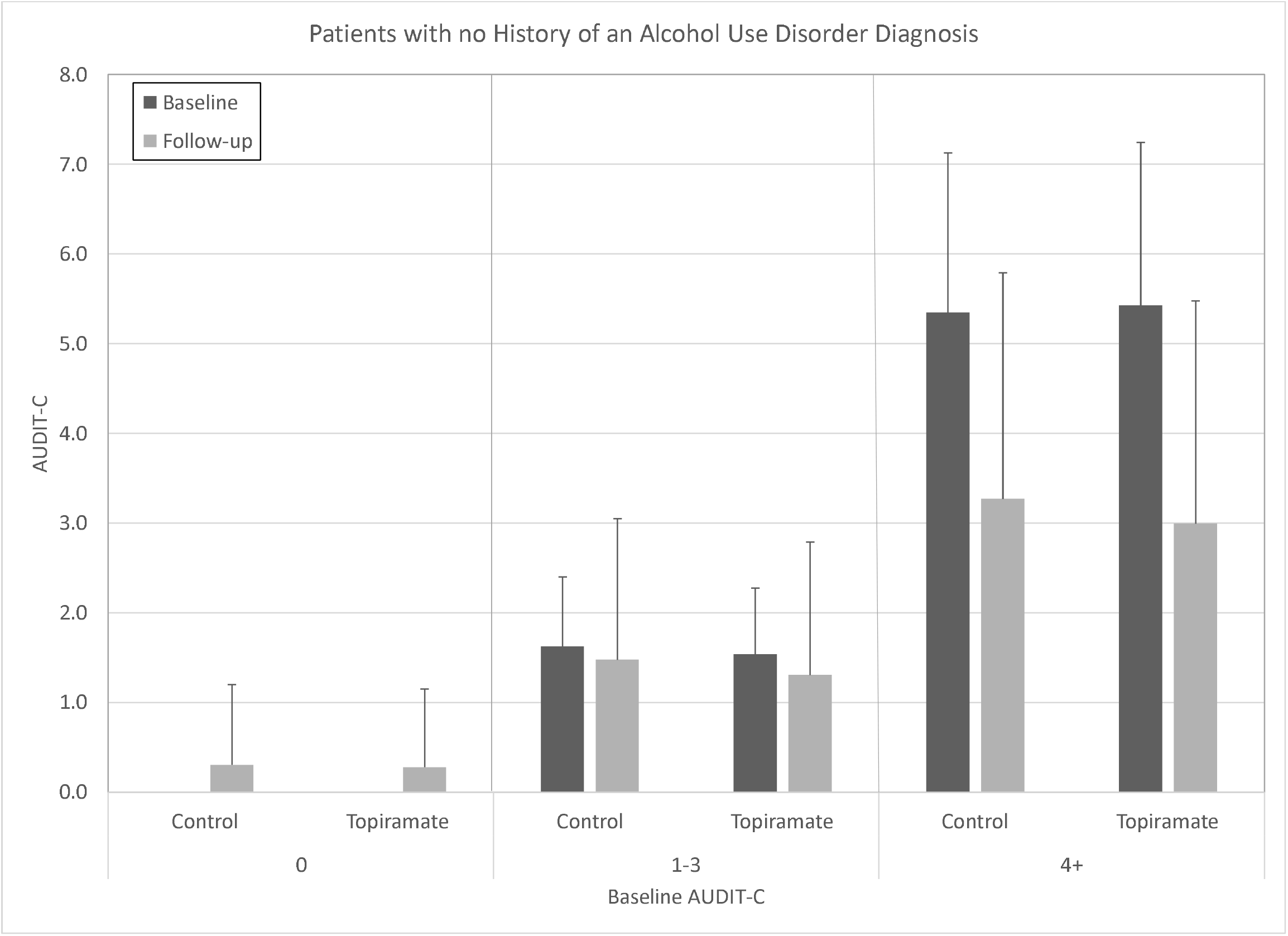
Baseline and Follow-up AUDIT-C Scores by Baseline AUDIT-C Score Group Among Patients with No History of an Alcohol Use Disorder Diagnosis.

Demographic and clinical characteristics at baseline for the final matched sample are presented in Table 1 and Supplementary Table 1. Among AUD-hx-pos patients, the mean age was 52.2 yr. (SD=12.0) and 5,254 (88.8%) were male, 4,314 (72.9%) were non-Hispanic white and 1,127 (19.0%) were non-Hispanic black. Within this subsample, only the number of neurology clinic visits (SMD=0.11) and number of prescriptions (SMD=0.65) were unbalanced between the exposure groups. Among AUD-hx-neg patients, the mean age was 53.6 yr (SD=14.7) and 18,649 (79.0%) were male, 18,168 (76.94%) were non-Hispanic white and 3,504 (14.8%) were non-Hispanic black. For AUD-hx-neg patients, number of visits to neurology clinics (SMD=0.13) and number of prescriptions (SMD=0.70) remained unbalanced between the exposure groups.

**Table 1:** Demographic and Clinical Measures of Unexposed Control and Topiramate Groups by History of Alcohol Use Disorder

|  | History of Alcohol Use Disorder<br>(n=5,918) |  |  |  |  | No History of Alcohol Use Disorder<br>(n=23,614) |  |  |  |  |
| --- | --- | --- | --- | --- | --- | --- | --- | --- | --- | --- |
|  | Topiramate (n=1,738) |  | Control (n=4,180) |  | Statistic | Topiramate (n=6,324) |  | Control (n=17,290) |  | Statistic |
| Age (M, SD) | 51.81 | 11.38 | 52.40 | 11.38 | -0.02 | 53.67 | 14.32 | 53.54 | 14.78 | 0.04 |
| Gender (n, %) |  |  |  |  | 0.00 |  |  |  |  | 0.01 |
| Female | 196 | 11.28 | 468 | 11.28 |  | 1,267 | 20.03 | 3,698 | 21.39 |  |
| Male | 1,542 | 88.72 | 3,712 | 88.72 |  | 5,057 | 79.97 | 1,3592 | 78.61 |  |
| Race (n, %) |  |  |  |  | 0.03 |  |  |  |  | 0.01 |
| White | 1,350 | 77.68 | 3,248 | 77.68 |  | 5,216 | 82.48 | 14,079 | 81.43 |  |
| Black or African American | 336 | 19.33 | 803 | 19.33 |  | 915 | 14.47 | 2,638 | 15.26 |  |
| American Indian/Alaskan Native | 15 | 0.86 | 24 | 0.86 |  | 43 | 0.68 | 127 | 0.73 |  |
| Asian | 11 | 0.63 | 21 | 0.63 |  | 37 | 0.59 | 103 | 0.60 |  |
| Native Hawaiian or Other Pacific Islander | 9 | 0.52 | 35 | 0.52 |  | 48 | 0.76 | 146 | 0.84 |  |
| Multiple Ancestries | 17 | 0.98 | 49 | 0.98 |  | 65 | 1.03 | 197 | 1.14 |  |
| Hispanic ethnicity | 76 | 4.37 | 234 | 5.60 | -0.03 | 240 | 3.80 | 1,103 | 5.86 | -0.04 |
| Body Mass Index (M, SD) | 31.01 | 6.77 | 30.83 | 6.77 | 0.01 | 32.00 | 6.73 | 31.83 | 7.03 | 0.01 |
| Smoker (past year) (n, %) | 1,155 | 66.46 | 2,755 | 66.46 | 0.01 | 2,302 | 36.4 | 6,304 | 36.46 | 0.00 |
| **# Prescriptions (M, SD)** | **82.7** | **75.34** | **27.07** | **53.29** | **0.65** | **72.64** | **64.88** | **22.32** | **43.89** | **0.70** |
| NSAIDs (n, %) | 316 | 18.18 | 735 | 18.18 | 0.01 | 957 | 15.13 | 2,441 | 14.12 | 0.01 |
| Opioids (n, %) | 441 | 25.37 | 1,011 | 25.37 | 0.01 | 1267 | 20.03 | 3,446 | 19.93 | 0.00 |
| Muscle relaxants (n, %) | 146 | 8.4 | 345 | 8.4 | 0.00 | 442 | 6.99 | 1,180 | 6.82 | 0.00 |
| Antidepressants (n, %) | 933 | 53.68 | 2,288 | 53.68 | -0.01 | 2,489 | 39.36 | 6,519 | 37.70 | 0.02 |
| # Hospitalizations (M, SD) | 0.54 | 1.17 | 0.48 | 1.17 | 0.05 | 0.21 | 0.63 | 0.17 | 0.56 | 0.04 |
| # Days hospitalized (M, SD) | 10.89 | 35.35 | 9.97 | 35.35 | 0.03 | 2.05 | 10.77 | 1.68 | 10.60 | 0.03 |
| # Outpatient Visits (M, SD) | 392.82 | 426.22 | 387.99 | 426.22 | 0.01 | 197.22 | 267.23 | 185.94 | 241.13 | 0.02 |
| **# Neurology Clinic Visits (M, SD)** | **0.38** | **0.86** | **0.24** | **0.75** | **0.11** | **0.40** | **0.87** | **0.24** | **0.85** | **0.13** |
| # Pain Clinic Visits (M, SD) | 0.20 | 0.91 | 0.17 | 1.14 | 0.01 | 0.22 | 1.10 | 0.13 | 0.95 | 0.06 |
| # Medical Diagnoses (M, SD) | 1.23 | 1.32 | 1.23 | 1.32 | 0.00 | 1.28 | 1.38 | 1.24 | 1.33 | 0.03 |
| # Psychiatric Diagnoses (M, SD) | 2.01 | 1.11 | 2.01 | 1.11 | -0.01 | 1.14 | 1.05 | 1.10 | 1.06 | 0.01 |
| # SUD Diagnoses (M, SD) | 0.49 | 0.56 | 0.50 | 0.56 | -0.01 | 0.26 | 0.44 | 0.26 | 0.44 | 0.00 |

M=mean; SD=standard deviation; NSAIDs=non-steroidal anti-inflammatory drugs; SUDs=substance use disorders. Statistics are phi for categorical values and SMD for continuous variables. \*Bolded values are significant at $\phi > 0.05$ or $SMD > 0.1$ . Note: The outpatient visit count comprises any visit, at any clinic, prior to the index date, covering a patient's entire medical history, as captured in the electronic health record.

### Baseline and Follow-up AUDIT-C Score

Average AUDIT-C scores at baseline and follow-up for AUD-hx-pos and AUD-hx-neg patients in the final matched sample are presented in Tables 2 and 3, respectively, which include additional statistics from the comparisons described in the text. Among AUD-hx-pos patients (Table 2), there were no significant differences in average AUDIT-C scores between exposure groups at baseline (2.44 for topiramate users versus 2.33 for controls; *P* = 0.24) or follow-up (1.81 versus 1.81; *P* = 0.98). Among AUD-hx-neg patients (Table 3), although average AUDIT-C score did not differ significantly between exposure groups at baseline (0.97 versus 1.01; *P* = 0.11), the topiramate group had a significantly lower average AUDIT-C score than the control group at follow-up (0.86 versus 0.96; *P* < 0.0001).

**Table 2.** Baseline AUDIT-C Scores, Average Daily Topiramate Dosage, and Current Alcohol Use Disorder Diagnosis Among Veterans with a History of an Alcohol Use Disorder Diagnosis

|  |  |  | Baseline |  |  |  |  | Follow-up |  |  |  |  | DiD |  |
| --- | --- | --- | --- | --- | --- | --- | --- | --- | --- | --- | --- | --- | --- | --- |
|  | Topiramate<br>[N (%)] | Control<br>[N (%)] | Topiramate<br>Mean (SD)<br>AUDIT-C<br>Score<br>(N=1,738) | Control<br>Mean (SD)<br>AUDIT-C<br>Score<br>(N=4,180) | t-<br>statistic | df | P-value | Topiramate<br>Mean (SD)<br>AUDIT-C<br>Score<br>(N=1,738) | Control<br>Mean (SD)<br>AUDIT-C<br>Score<br>(N=4,180) | t-<br>statistic | df | P-value | Estimate | P-value |
|  |  |  | 2.44 (3.50) | 2.33 (3.38) | -1.17 | 5,916 | 0.24 | 1.81 (3.01) | 1.81 (2.87) | -0.02 | 3,116 | 0.98 | -0.11 | <.0001 |
| **AUDIT-C Subgroup** |  |  |  |  |  |  |  |  |  |  |  |  |  |  |
| 0 | 869 (50.0) | 2,119 (50.7) | 0.00 | 0.00 |  |  |  | 0.61 (1.87) | 0.70 (1.89) | 1.22 | 2,986 | 0.22 | -0.09 | 0.38 |
| 1-3 | 401 (23.1) | 963 (23.0) | 1.83 (0.83) | 1.82 (0.82) | -0.26 | 1,362 | 0.79 | 2.00 (2.61) | 1.95 (2.36) | -0.34 | 685 | 0.73 | 0.04 | 0.81 |
| 4-7 | 247 (14.2) | 604 (14.5) | 5.16 (1.11) | 5.07 (1.06) | -1.07 | 849 | 0.29 | 3.39 (3.17) | 3.44 (3.08) | 0.18 | 849 | 0.86 | -0.13 | 0.52 |
| 8+ | 221 (12.7) | 494 (11.8) | 10.14 (1.49) | 9.98 (1.49) | -1.35 | 713 | 0.18 | 4.44 (4.31) | 4.31 (4.15) | -0.37 | 713 | 0.71 | -0.04 | 0.87 |
| **Topiramate Dosage Subgroup (mg/day)** |  |  |  |  |  |  |  |  |  |  |  |  |  |  |
| Low<br>(≤100) | 927 (53.3) | 2,169 (51.9) | 2.35 (3.35) | 2.23 (3.27) | -0.94 | 3,094 | 0.34 | 1.76 (2.92) | 1.81 (2.83) | 0.43 | 3,094 | 0.66 | -0.20 | 0.24 |
| Medium<br>(>100,<br>≤150 mg) | 262 (15.1) | 585 (14.0) | 2.70 (3.82) | 2.56 (3.49) | -0.53 | 845 | 0.60 | 2.13 (3.38) | 1.82 (2.94) | -1.31 | 4,445 | 0.19 | -0.05 | 0.75 |
| High<br>(>150 mg) | 549 (31.6) | 1,426 (34.1) | 2.48 (3.60) | 2.39 (3.47) | -0.51 | 1973 | 0.61 | 1.74 (2.97) | 1.81 (2.91) | 0.43 | 1,973 | 0.66 | -0.10 | 0.55 |
| **Current Alcohol Use Disorder** |  |  |  |  |  |  |  |  |  |  |  |  |  |  |
| No | 875 (50.3) | 2,111 (50.5) | 1.38 (2.39) | 1.39 (2.38) | 0.04 | 2,984 | 0.97 | 1.19 (2.20) | 1.35 (2.31) | 1.83 | 2,984 | 0.068 | -0.16 | 0.22 |
| Yes | 863 (49.7) | 2,069 (49.5) | 3.52 (4.08) | 3.29 (3.93) | -1.41 | 2,930 | 0.16 | 2.45 (3.54) | 2.28 (3.28) | -1.21 | 1,509 | 0.23 | -0.06 | 0.67 |

SD=standard deviation; AUDIT-C=Alcohol Use Disorders Identification Test-Consumption; df=degrees of freedom; DiD=difference-in-difference score.

**Table 3.** Baseline and Follow-up AUDIT-C Scores, Average Daily Topiramate Dosage Among Veterans with No History of an Alcohol Use Disorder Diagnosis

|  |  |  | Baseline |  |  |  |  | Follow-up |  |  |  |  | DiD |  |
| --- | --- | --- | --- | --- | --- | --- | --- | --- | --- | --- | --- | --- | --- | --- |
|  | Topiramate<br>[N (%)] | Control<br>[N (%)] | Topiramate<br>Mean (SD)<br>AUDIT-C<br>Score<br>(N=6,324) | Control<br>Mean (SD)<br>AUDIT-C<br>Score<br>(N=17,290) | t-<br>statistic | df | P-value | Topiramate<br>Mean (SD)<br>AUDIT-C<br>Score<br>(N=6,324) | Control<br>Mean (SD)<br>AUDIT-C<br>Score<br>(N=17,290) | t-<br>statistic | df | P-value | Estimate | P-value |
|  |  |  | 0.97 (1.62) | 1.01 (1.63) | 1.61 | 23,612 | 0.11 | 0.86 (1.52) | 0.96 (1.61) | 4.71 | 11,870 | <0.0001 | -0.07 | 0.0002 |
| **AUDIT-C Subgroup** |  |  |  |  |  |  |  |  |  |  |  |  |  |  |
| 0 | 3,591 (56.8) | 9,703 (56.1) | 0.00 | 0.00 |  |  |  | 0.28 (0.87) | 0.31 (0.90) | 1.56 | 6,580 | 0.119 |  |  |
| 1-3 | 2,242 (35.4) | 6,230 (36.0) | 1.54 (0.74) | 1.63 (0.78) | 4.68 | 4,156 | <0.0001 | 1.31 (1.48) | 1.48 (1.57) | 4.63 | 4,174 | <0.0001 | -0.09 | 0.011 |
| 4+ | 491 (7.8) | 1,357 (7.8) | 5.43 (1.82) | 5.35 (1.78) | -0.82 | 1,846 | 0.41 | 3.00 (2.48) | 3.27 (2.52) | 2.08 | 1846 | 0.038 | -0.35 | <0.0001 |
| **Topiramate Dosage Subgroup (mg/day)** |  |  |  |  |  |  |  |  |  |  |  |  |  |  |
| Low<br>(≤100) | 3,598 (56.9) | 9,650 (55.9) | 1.01 (1.67) | 1.04 (1.66) | 0.92 | 13,246 | 0.36 | 0.90 (2.59) | 0.98 (1.62) | 2.59 | 6,677 | 0.0095 | -0.05 | 0.09 |
| Medium<br>(>100,<br>≤150 mg) | 864 (13.7) | 2,329 (13.5) | 0.99 (1.54) | 1.10 (1.66) | 1.74 | 1,653 | 0.08 | 0.92 (1.20) | 1.00 (1.64) | 1.2 | 3,191 | 0.2297 | 0.03 | 0.60 |
| High<br>(>150 mg) | 1,862 (29.4) | 5,311 (30.7) | 0.88 (1.57) | 0.91 (1.54) | 0.67 | 7,171 | 0.50 | 0.73 (1.39) | 0.91 (1.58) | 4.51 | 3,655 | <0.0001 | -0.148 | 0.0003 |

SD=standard deviation; AUDIT-C=Alcohol Use Disorders Identification Test-Consumption; df=degrees of freedom; DiD=difference-in-difference score.

Among AUD-hx-pos patients, the estimated change in AUDIT-C score was -0.63 [SE=0.08, 95%CI=(−0.79,-0.47), *P* < 0.0001] for topiramate-treated patients and -0.52 [SE=0.05, 95%CI=(−0.62, -0.42), *P* < 0.0001] for matched controls, resulting in a DiD score of -0.11 [SE=0.10, 95%CI= (−0.30,0.08), *P* = 0.24]. There were no significant effects of baseline AUDIT-C score (F(2,5910)=0.21, *P* = 0.89), average total daily topiramate dosage (F(2,5912)=0.79, *P* = 0.45), or a current diagnosis of AUD (F(1,5914)=0.31, *P* =0.58) on the average AUDIT-C DiD score in AUD-hx-pos patients.

Among AUD-hx-neg patients, the estimated change over time in AUDIT-C score was -0.11 [SE=0.02, 95%CI=(−0.15, -0.07), *P* < 0.0001] for topiramate-treated patients and -0.04 [SE=0.01, 95%CI=(−0.07, -0.02, *P* = 0.0002] for matched controls, a significant difference by exposure group [DiD= -0.07, SE=0.02, 95%CI=(−0.11, -0.03), *P* = 0.002). Baseline AUDIT-C score was associated with the DiD score (F(2,24000)=9.06, *P* = 0.0001), such that higher baseline AUDIT-C scores were associated with a greater DiD score favoring the topiramate group. Although the effect of topiramate was significant among patients with baseline AUDIT-C scores of 1-3 (DiD=-0.09, SE=0.03, 95%CI=(−0.15, -0.02), *P* = 0.011), it was substantially larger among those with baseline AUDIT-C scores of 4+ (DiD=-0.35, SE=0.07, 95%CI=(−0.49, -0.21), *P* < 0.0001). There was also a significant effect of average daily topiramate dosage on the DiD score in AUD-hx-neg patients (F(2, 24,000)=3.43, *P* = 0.03). At a high average dosage (i.e., >150 mg/day), the estimated change over time in AUDIT-C score was -0.15 [SE=0.04, 95%CI=(−0.22, -0.08), *P* < 0.0001] for topiramate-treated patients and -0.002 [SE=0.02, 95%CI=(−0.04, 0.04), *P* = 0.91] for matched controls, a significant DiD of -0.148 [beta= -0.15, SE=0.04, 95%CI=(−0.23, -0.07), *P* = 0.0003]. The effects of topiramate were not significant at the two lower dosage levels [<100 mg/day: beta= -0.05, SE=0.03, 95%CI=(−0.11, 0.01), *P* = 0.09] and 100-150 mg/day: beta=0.03, SE=0.06, 95%CI=(−0.09, 0.15), *P* = 0.60].

A sensitivity analysis in which we excluded patients who reported no drinking at an assessment closest in time to the baseline (AUDIT-C=0) yielded similar results.

## DISCUSSION

Using EHR data from the largest integrated healthcare system in the United States, we found that both AUD-hx-pos and AUD-hx-neg individuals reported significantly decreased drinking over time. Among AUD-hx-pos individuals, the change over time was similar between the topiramate-exposed and unexposed groups, even when considering individuals with a current AUD diagnosis. Among individuals with no history of AUD, topiramate was associated with reduced drinking. This small effect was most evident among patients with higher baseline drinking levels. Patients with a baseline AUDIT-C score of 1-3 had a DiD score of -0.09, while those with a baseline AUDIT-C score of 4+ had a DiD score of -0.35. A subanalysis also revealed that the effect of topiramate was present only among individuals receiving an average daily dosage of >150 mg/day, where the DiD score was -0.148. The effects on AUDIT-C score, although statistically significant, represent a small reduction in one or more of the 3 AUDIT-C items: frequency of drinking, quantity of drinking, or frequency of intoxication. For example, a DiD score of -0.35 in individuals with an AUDIT-C score of 4+ could reflect a reduction in the frequency of drinking, but would not be large enough to go from 2-4 times per month (which is scored 2) to monthly or less (which is scored 1), as that would be a DiD score of -1.0.

The absence of an effect of topiramate in AUD-hx-pos patients, particularly those with a current AUD diagnosis, contrasts with the findings from multiple randomized controlled trials (RCTs) of topiramate for treating AUD, which showed a robust reduction in heavy drinking (6, 26). Although the AUDIT-C is validated and widely used as a screening instrument, the quality of alcohol screening (and thus the accuracy of the alcohol consumption estimate) cannot be assured without careful monitoring and quality control (27). An observational study of AUDIT-C screening showed that most screening was conducted verbally and included non-verbatim screening and making inferences, assumptions, and/or suggestions to input responses and adaptations of screening questions to enhance patient comfort (28). These non-standard methods could reduce the accuracy of the information obtained. Thus, topiramate’s effect on drinking could have been seen only in AUD-hx-neg patients because they may provide more accurate self-reports of drinking than patients with AUD, particularly those not seeking alcohol treatment.

The findings reported here also highlight differences between motivated volunteers in an RCT who seek to reduce or stop drinking and patients in clinical care who may not have sought to change their drinking behavior. Thus, these findings could indicate that although topiramate helps people reduce their drinking if they seek to do so, in the absence of such motivation (and the psychosocial support that often accompanies study medication in an RCT for AUD) topiramate does not substantially reduce drinking.

In the AUD-hx-neg group, the dose-related effect of topiramate is consistent with a pharmacological effect of the medication on drinking. However, the small effect limits its clinical significance. Larger medication effects have been reported in prior EHR-based studies that used propensity-score matching to examine the effects of gabapentin (29) and spironolactone (30) prescribed for any indication on alcohol consumption.

Rentsch et al. (29) compared patients from the Veterans Aging Cohort Study who were prescribed gabapentin with up to 5 matched, unexposed patients. They found that gabapentin-exposed patients showed a decrease in AUDIT-C score that was 0.39 points greater than in unexposed patients. The effect of gabapentin was nearly twice that (DiD score = 0.77) among patients with a history of AUD who received ≥1,500 mg/d of the medication. Effects of gabapentin were not significant among AUD-hx-pos individuals who received a lower dosage of gabapentin or had a baseline AUDIT-C score ≥4, as well as AUD-hx-neg individuals.

Palzes et al. (30) used EHR data from Kaiser Permanente Northern California, a large integrated healthcare system, to examine changes in weekly alcohol use among 523 spironolactone-treated and 2,305 untreated adults. They found that treated spironolactone-treated patients reduced their alcohol use by 0.76 drinks/week more than untreated patients. Follow-up analyses revealed that the effect of spironolactone was seen only among patients who at baseline were drinking >7 drinks/week. Further, a higher spironolactone dosage was associated with a significantly greater DiD score.

Notably, there is considerably greater evidence from clinical trials that topiramate consistently reduces alcohol consumption (6, 26) than is true for either gabapentin (31) or spironolactone (for which there are no published RCTs in AUD). Thus, efforts are needed to identify key factors that influence the validity of EHR data for evaluating medications to treat AUD, such as self-reported drinking behavior. This could help to increase the correspondence between effects observed in EHR data and those from RCTs, the gold standard for evaluating efficacy. This would help to define the utility of large, extant databases in AUD medications development.

Limitations of this study include a reliance on self-reported drinking behavior, which as suggested above, could have contributed to the lack of a medication effect in the AUD-hx-pos group and possibly an underestimation of its effect in AUD-hx-neg individuals. Use of a biological measure of alcohol consumption, such as phosphatidylethanol (PEth) (32, 33), could help to increase the accuracy of the outcome measure in analyses such as this. We chose to limit the study to the period prior to 2015, when ICD-9 was replaced with ICD-10, to ensure consistency in disease coding. However, the use of topiramate to treat AUD has increased in recent years, so that this may have reduced the number of individuals in the study sample with current AUD who received topiramate treatment. This could have contributed to the lack of an effect of topiramate treatment in patients with current AUD. Because PEth is not a routine laboratory test in the VA, it was not available for most patients in the study. Other, routine laboratory tests, such as gamma-glutamyl transferase, are less sensitive and specific than PEth (33) and results from these tests are also not consistently available in the VA EHR. The EHR, although a useful source of diagnostic information, does not cover a patient’s entire lifetime, so it likely resulted in an underestimation of the prevalence of a history of AUD. Given the chronic nature of AUD and the substantial efforts of the VA healthcare system to identify and treat AUD, the impact of this undercounting is likely to be small. Because the patient population studied was predominantly male and comprised exclusively of veterans who receive care in the VA healthcare system, our ability to generalize the findings to women, veterans who receive care outside the VA healthcare system, and non-veterans is limited. Further, the age of our sample (∼52 years) differs substantially from individuals with AUD in the population, whose mean age at first AUD treatment has been reported to be 29.4 years (34). Two factors mitigate the impact of this difference, however. First, the age at which AUD-hx-pos veterans in our study received topiramate is likely later than when they may first have been treated for AUD. Second, the mean age of participants in some RCTs of topiramate for treating AUD is close to that of the patients in our sample. For example, in the largest multi-center trial of topiramate for treating AUD the mean age of participants was 47.3 years (35).

The study’s strengths include the large sample, which provides adequate statistical power to test small effects. Propensity-score matching permitted a balanced comparison of exposed and unexposed patients and lends confidence in the validity of the findings, which suggest that prescribing topiramate to individuals without a history of AUD for a variety of indications can modestly reduce drinking. Recent epidemiological findings show that, even at low levels of alcohol consumption there is a discernible increase in all-cause mortality, particularly deaths due to cancer (36). Thus, even a small decrease in alcohol consumption could have beneficial effects on public health. Further research is needed to ascertain whether the use of data from the AUDIT-C accurately estimates the reduction in drinking associated with topiramate treatment and, given the observed effects, which patient characteristics can be used to identify patients most likely to reduce their drinking.

## Data Availability

All data produced in the present study are available upon reasonable request to the authors, within the constraints of the Department of Veterans Affairs regulations.

