## Supplemental Files for "Association of Topiramate Prescribed for any Indication with Reduced Alcohol Consumption in Electronic Health Record Data"

|  | History of Alcohol Use Disorder | | | | | | | No History of Alcohol Use Disorder | | | | | | |
| --- | --- | --- | --- | --- | --- | --- | --- | --- | --- | --- | --- | --- | --- | --- |
|  | Control  (n=18,610) | | Topiramate (n=3,722) | |  | | | Control  (n=64,565) | | Topiramate (n=12,913) | |  | | |
|  | n | % | n | % | Statistic* | df | P-value | n | % | n | % | Statistic* | df | P-value |
| Psychiatric Diagnoses |  |  |  |  |  |  |  |  |  |  |  |  |  |  |
| PTSD | 10,302 | 55.36 | 2,047 | 55.00 | 0.1626 | 1 | 0.6867 | 22,039 | 34.13 | 4,327 | 33.51 | 1.8767 | 1 | 0.1707 |
| Depressive Disorder | 15,094 | 81.11 | 2,995 | 80.47 | 0.8241 | 1 | 0.3640 | 34,805 | 53.91 | 6,850 | 53.05 | 3.1986 | 1 | 0.0737 |
| Bipolar disorder | 6,896 | 37.06 | 1,378 | 37.02 | 0.0014 | 1 | 0.9703 | 9,046 | 14.01 | 1,777 | 13.76 | 0.5568 | 1 | 0.4556 |
| Schizophrenia | 1,673 | 8.99 | 324 | 8.70 | 0.3089 | 1 | 0.5783 | 2,013 | 3.12 | 384 | 2.97 | 0.7447 | 1 | 0.3882 |
| Schizoaffective disorder | 1,504 | 8.08 | 299 | 8.03 | 0.0098 | 1 | 0.9212 | 1,753 | 2.72 | 343 | 2.66 | 0.1416 | 1 | 0.7067 |
| Generalized Anxiety Disorder | 2,037 | 10.95 | 410 | 11.02 | 0.0155 | 1 | 0.9009 | 3,923 | 6.08 | 782 | 6.06 | 0.0076 | 1 | 0.9303 |
| Social Anxiety Disorder | 236 | 1.27 | 44 | 1.18 | 0.1852 | 1 | 0.6670 | 249 | 0.39 | 50 | 0.39 | 0.0007 | 1 | 0.9793 |
| Panic Disorder | 1,263 | 6.79 | 233 | 6.26 | 1.3761 | 1 | 0.2408 | 2,425 | 3.76 | 481 | 3.72 | 0.0286 | 1 | 0.8657 |
| Obsessive-Compulsive Disorder | 433 | 2.33 | 82 | 2.20 | 0.2103 | 1 | 0.6465 | 710 | 1.10 | 146 | 1.13 | 0.0945 | 1 | 0.7585 |
| Agoraphobia | 568 | 3.05 | 114 | 3.06 | 0.0012 | 1 | 0.9723 | 1,022 | 1.58 | 192 | 1.49 | 0.6434 | 1 | 0.4225 |
| Specific phobia | 34 | 0.18 | 6 | 0.16 | 0.0801 | 1 | 0.7771 | 116 | 0.18 | 19 | 0.15 | 0.6545 | 1 | 0.4185 |
| Hypochondria | 7 | 0.04 | 1 | 0.03 | 0.1000 | 1 | 0.7518 | 14 | 0.02 | 5 | 0.04 | 1.2740 | 1 | 0.2590 |
| Substance Use Disorders |  |  |  |  |  |  |  |  |  |  |  |  |  |  |
| Tobacco | 11,872 | 63.79 | 2,370 | 63.68 | 0.0188 | 1 | 0.8911 | 18,409 | 28.51 | 3,621 | 28.04 | 1.1723 | 1 | 0.2789 |
| Cocaine | 4,467 | 24.00 | 922 | 24.77 | 1.0003 | 1 | 0.3172 | 660 | 1.02 | 147 | 1.14 | 1.4087 | 1 | 0.2353 |
| Alcohol | 18,610 | 100 | 3,722 | 100 | -- | -- | -- | 505 | 0.78 | 99 | 0.77 | 0.0334 | 1 | 0.8550 |
| Opioids | 2,333 | 12.54 | 472 | 12.68 | 0.0594 | 1 | 0.8074 | 1,082 | 1.68 | 217 | 1.68 | 0.0014 | 1 | 0.9701 |
| Stimulants | 856 | 4.60 | 181 | 4.86 | 0.4856 | 1 | 0.4859 | 224 | 0.35 | 44 | 0.34 | 0.0120 | 1 | 0.9128 |
| Marijuana | 4,000 | 21.49 | 818 | 21.98 | 0.4287 | 1 | 0.5126 | 1,290 | 2.00 | 259 | 2.01 | 0.0033 | 1 | 0.9542 |
| Sedatives | 739 | 3.97 | 149 | 4.00 | 0.0084 | 1 | 0.9268 | 255 | 0.39 | 50 | 0.39 | 0.0165 | 1 | 0.8979 |
| Hallucinogen | 88 | 0.47 | 17 | 0.46 | 0.0172 | 1 | 0.8956 | 9 | 0.01 | 3 | 0.02 | 0.6001 | 1 | 0.4385 |
| Other drug use | 5,092 | 27.36 | 1,034 | 27.78 | 0.2737 | 1 | 0.6009 | 1,019 | 1.58 | 218 | 1.69 | 0.8283 | 1 | 0.3628 |
| Combination drug use | 3,721 | 19.99 | 776 | 20.85 | 1.4078 | 1 | 0.2354 | 458 | 0.71 | 92 | 0.71 | 0.0015 | 1 | 0.9695 |
| Unspecified drug use | 2,656 | 14.27 | 542 | 14.56 | 0.2128 | 1 | 0.6445 | 355 | 0.55 | 70 | 0.54 | 0.0118 | 1 | 0.9134 |

Supplementary Table 1: Prevalence of Psychiatric and Substance Use Disorder Diagnoses of Unexposed Control and Topiramate Groups by History of Alcohol Use Disorder

*Chi-square statistic
